# Computer Vision-Based Retrieval of Steps and Errors in Laparoscopic Cholecystectomy

**DOI:** 10.64898/2026.01.15.25343153

**Authors:** Elaine Sui, Charlotte Egeland, Xiaohan Wang, Alfred Song, Rui Li, Joshua Villarreal, Anita Rau, Josiah Aklilu, Alan Brown, Shelly Goel, Brian Sutjiadi, Roger Bohn, Eric Sorenson, Vanessa Palter, Teodor Grantcharov, Jeffrey Jopling, Serena Yeung-Levy

## Abstract

Traditional surgical training relies heavily on hands-on experiences gained through relatively infrequent procedures during apprenticeships. Recently, postoperative review has become a valuable supplement to this model, offering learning opportunities outside the operating room. However, its adoption remains limited due to its inefficiencies. In this study, we developed a Computer Vision-based system designed to efficiently navigate and retrieve critical segments from laparoscopic cholecystectomy videos. Trained on a manually annotated dataset of 683 videos from six different sources, our model is taught to identify five distinct surgical steps and three common intraoperative errors: bleeding, bile spillage, and thermal injury. The resulting models demonstrated strong generalizability in identifying steps to unseen data from new hospitals. Large-scale analysis further revealed correlations between model-predicted error events and surgical step, disease severity, and surgical skill that align with established clinical knowledge. By facilitating the rapid retrieval of clinically relevant events, this tool enhances postoperative review practical, ultimately expediting surgical training.

## INTRODUCTION

The traditional apprenticeship model of surgical education is evolving into a competency-based, data-driven approach. While surgical video review is increasingly used in U.S. training programs, its educational potential remains underutilized^1,2^. Studies have shown that video-coaching can improve technical skills^3^ and that postoperative video review enhances both surgical skill acquisition^4^ and the learning experience^5^. Furthermore, there is an association between higher technical skills and patient outcomes^6^. Yet, barriers such as time constraints and challenges with video upload and editing often hinder surgeons in their efforts to adopt video review into their daily practice^4,7^. The ability to pinpoint key moments within a procedure could greatly enhance the feasibility and value of postoperative video review, as trainees can extract clips of interest for self-study and to present to attendings for more targeted discussion.

Recent advances in Computer Vision (CV) have opened new possibilities for enhancing surgical training programs and assisting postoperative video review. Technical progress in vision foundation models^8,9^, combined with an increasing number of publicly available surgical datasets^10–14^, has significantly expanded the potential for CV in healthcare applications^10–12,15–17^ Despite these advances, few studies have focused on localizing key moments within a procedure, which is critical for efficient postoperative video review.

We hypothesize that CV can be leveraged to build an efficient system for navigating to key moments within surgical procedure videos. In this study, we developed an automated system designed to efficiently navigate to and retrieve specific segments of interest from full-length laparoscopic cholecystectomy (LC) videos. We applied this system across multiple temporal scales, focusing on specific surgical steps and errors. Unlike prior approaches that have examined surgical steps^10,18^ and errors^19^ independently, our work is the first to integrate both, enabling a more comprehensive analysis. Compared to previous research on error retrieval^19^, our system provides more meaningful insights by tailoring error definitions specifically to LC, capturing instances that are clinically relevant for skill assessment and training. At the core of our approach is an ontology that organizes this multi-scale breakdown into coarser steps and fine-grained errors. We developed a CV-based temporal model to retrieve relevant segments from a full-length video. Since models face unique challenges when processing videos of varying lengths and temporal resolutions, we incorporated architectural modifications and utilized specific training and inference strategies to address these issues. Furthermore, this work demonstrates the practical applicability of our framework in a large-scale cohort, showing how our step model generalizes to LC videos from new hospitals and how findings from our error model aligns with known clinical realities.

## METHODS

### Dataset

We assembled a dataset of LC videos from three medical institutions as well as one private company, Surgical Safety Technologies Inc. (SST), New York, NY, USA. Videos were collected from two of the medical institutions using the Black Box Explorer™ platform. This dataset was further enriched by incorporating videos available from two public datasets (Cholec80^10^, Hei-Chole^14^). The videos were originally recorded from procedures in Europe and the US. No robotic assisted cases were included.

Surgeon experience level data was available for a subset of cases from Institution 1, and for all cases from Institutions 2 and 3. To protect patient confidentiality, all videos used to train and test the models were de-identified. Videos from Institution 1, which were directly acquired, were manually de-identified by blacking out portions where the camera was out of the body. Videos from SST, Institution 2 and Institution 3, which were downloaded from Black Box Explorer™, were automatically de-identified using a CV algorithm integrated in the Black Box Explorer™ platform prior to transmission. The videos were then downsampled temporally to 10 frames per second (fps) and spatially to 480×854 pixels to speed up video processing and analysis.

### Ontology & Annotations

To train the CV models, we manually labeled the data using an ontology based on recent recommendations from the Society of American Gastrointestinal and Endoscopic Surgeons (SAGES) video annotation framework working group^20^. We concentrate on the Execution of Surgical Objectives phase, which we break down into specific steps. Comparable step ontologies have been developed previously based on the same framework^21–23^.

We focused on laparoscopic skills, developing an ontology of five intracorporeal steps: Exposure of the Gallbladder, Dissection of the Hepatocystic Triangle, Ligation & Division, Gallbladder Dissection, and Packaging & Removal (Supplementary Table 1). Following previous work^24,25^, we also defined surgical errors within these steps. Error instances are moments in the procedure where the surgeon deviates from optimal performance, and thus includes, but is not limited to, clinically serious adverse events. We concentrated on detecting bleeding, bile spillage, and thermal injury, as these are the most common events with significant impact on task progression. Bleeding is the active flow of blood from an injury whereas bile spillage is the active flow of bile caused by a tool-tissue interaction. A single injury can generate multiple error instances. For example, an untreated bleed may persist off-camera and be recorded as a new error event when the camera shifts focus back to the same bleed. The same applies to bile spillage. There was no differentiation between minor and major bleeding episodes, nor between perforations of the gallbladder, cystic duct, or common bile duct. Thermal injury is defined as a tool-tissue interaction that leads to a burn mark outside of the surgical bed. This includes, but is not limited to, burn marks on the liver or duodenum. A burn mark is counted only once as an error, regardless of its duration or camera re-focusing.

The ontology was developed by surgeons and surgical residents who have each performed over 150 laparoscopic cholecystectomies. Annotators were trained in-house by the ontology developers through ontology review, example cases, and pre-labelled video demonstrations.

Performance was tested using the same videos for comparison, and quality checks were conducted throughout the labelling process to ensure accuracy and consistency.

A 4-point disease severity score was labelled by a trained surgeon for all procedures where the initial state of the gallbladder was visible, ranging from normal-appearing to severely inflamed gallbladder^26^ (Supplementary Table 2).

All data was annotated using Labelbox^27^.

### Temporal Modelling for Navigation to Steps and Errors

In this study, we developed CV-based models to navigate and retrieve five surgical steps and three common errors from full-length videos. As shown in Figure 1, the models share a similar architecture of a per-frame image feature extractor followed by a temporal modelling layer to capture the interdependence between video frames. However, as there are different challenges in retrieving long, coarse-grained and short, fine-grained segments, the models are specifically tailored to these different tasks.

**Figure 1.**
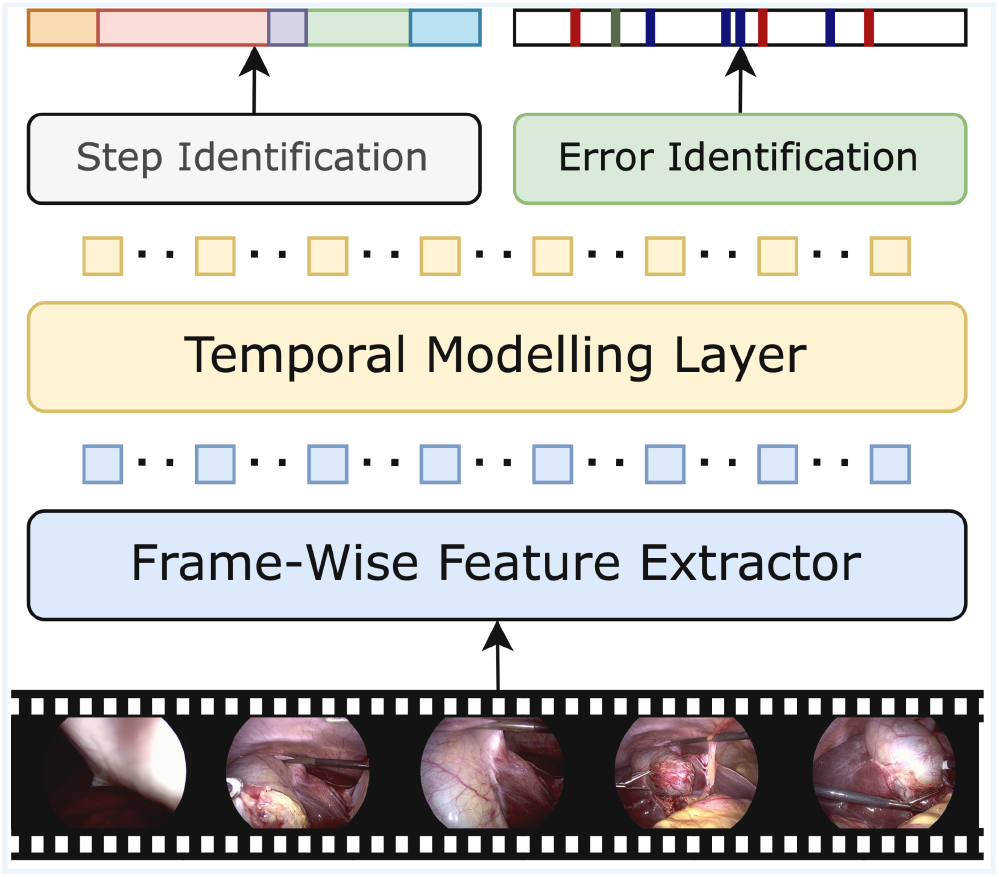
Computer-Vision Model Architecture. Diagram of Computer Vision-based method to identify surgical steps and errors in a full-length laparoscopic procedure.

For surgical step identification, the model outputs a step label for every frame of a full-length procedure at once (Supplementary Figure 1). As this is a coarse-grained task, we can simply extract pre-computed image features and only train the temporal and step classification layers on video frames sampled at 1 fps. In contrast, for surgical error identification, the model produces a binary label indicating whether a tissue injury occurred within a short 16-frame clip (Supplementary Figure 2). The error model is then applied in a sliding-window fashion over the entire full-length procedure to identify all predicted error instances (Supplementary Figure 3). As this is a fine-grained task, we train the entire model, including the feature extractor, at higher temporal resolutions up to 10 fps as opposed to the lower 1 fps for the step model.

To evaluate models that output framewise labels for full-length videos, such as for surgical step identification, we report standard metrics of framewise accuracy, precision, and recall. For models identifying multiple instances of an event, such as surgical errors, we evaluate the precision and recall of the identified instances. Specifically, for error identification, we use a loose precision and recall definition, considering a “hit” as any predicted error occurring within five seconds of the actual error. The five-second buffer accounts for the delayed visibility of tissue injuries that may initially be occluded by surgical tools. This allows time for the tools to move, revealing the injury the model detects.

We report these metrics averaged per video (video-level) and across all instances (instance-level). Additionally, given that many videos contain no instances of certain errors, we include video-level accuracy for error presence to assess the model’s ability to correctly identify videos as completely error-free.

To train and evaluate our Surgical Step Recognition model, we randomly split our dataset by procedure into 50% training, 10% validation, and 40% testing for each source, except for Cholec80, where the first 40 videos are used for training, the next 8 for validation and the last 32 for testing. To train and evaluate our Surgical Error Detection model, with the exception of Cholec80 where the previously described split is used, we randomly split our dataset by procedure into 70% training, 15% validation and 15% testing for each source such that videos in the training split for the step recognition model are a subset of the training split for error detection.

### Surgical Error Correlation Analysis

We applied our error detection model on the entire video dataset to identify correlations with known metadata using the Mann–Whitney U test^28^ and a Kruskal–Wallis test^29^ (Supplemental Content 5). We also compared these correlations with those derived from ground truth (GT) annotations (Supplementary Figure 4). Since GT annotations are only available for a smaller subset of our full dataset, a few correlations observed using model predictions are not as strongly observed when using GT annotations. However, these correlations are consistent with those derived from GT annotations either in a statistically significant manner or when observing the pattern of median values across groups, supporting the utility of our CV models. We analyzed error predictions stratified by surgical step, disease severity score (DSS), and surgeon experience level (attending vs. resident).

The study received ethical approval from the Stanford Institutional Review Board (IRB-57677).

## RESULTS

### Dataset

Our study dataset comprised 683 videos totaling over 650 hours from six data sources (Table 1). Videos were temporally annotated at 10 fps for surgical steps and errors based on our defined ontology, resulting in 2,186 annotated instances of bleeding from 285 videos, 1,239 instances of bile spillage from 683 videos, and 1,384 instances of thermal injury from 606 videos. Notably, no instances of common bile duct injury were observed in the dataset. Out of the 285 videos annotated for all error types, 6 (2.1%) had no error instances.

**Table 1.** Details of all data sources used in this work, including the video distribution across institutions, total video length, and when available, the number of videos performed by a surgical resident vs. an attending surgeon as the primary surgeon.

| **Dataset** | **Provider** | **Videos** | **Total Time (hours)** | **Performed by Surgical Resident** | **Performed by an Attending** |
| --- | --- | --- | --- | --- | --- |
| Cholec80 | University of Strasbourg | 80 | 51.25 | N/A | N/A |
| Hei-Chole | University of Heidelberg | 23 | 14.57 | N/A | N/A |
| SST | Surgical Safety Technologies | 222 | 278.68 | N/A | N/A |
| Institution 1 | Institution 1 | 275 | 207.48 | 119 | 153 |
| Institution 2 | Institution 2 | 39 | 47.30 | 39 | 0 |
| Institution 3 | Institution 3 | 44 | 54.80 | 44 | 0 |

Among the 355 videos annotated with surgical experience, 43.1% were performed by attendings, and 56.9% were performed by surgical residents as the primary operator. For the 512 videos annotated with disease severity, 38.5% were classified as DSS 1 (normal appearing gallbladder), 25.2% as DSS 2 (mildly abnormal appearing gallbladder), 30.4% as DSS 3 (moderately abnormal appearing gallbladder), and 5.9% as DSS 4 (severely inflamed or grossly abnormal appearing gallbladder).

### Surgical Step Recognition

We built a Computer Vision Surgical Step Recognition model to predict one of five surgical steps for each video frame. Across all videos, the model achieved an average framewise accuracy of 88.3%, precision of 83.1%, and recall of 86.6% (dataset-specific metrics found in Supplementary Table 3).

### Surgical Step Generalization Analysis

### Surgical Error Detection

We used our Surgical Error Detection models to localize bleeding, bile spillage, and thermal injury within each video. Table 2 describes the performance of our model to detect each instance of the three types of errors in full-length video procedures, achieving high video-level and instance-level recalls of up to 62.6% and 55.8%, respectively, and moderate video-level and instance-level precision of up to 28.7% and 21.7%, respectively (detailed error performance results are in Supplementary Table 4 and Supplementary Table 5). For postoperative review settings, high recall is prioritized over precision in order to ensure retrieval of an adequate number of true instances (Supplemental Content 4). On average, our model predicts 30.8 bleeding events, 3.2 bile spillage events, and 5.0 thermal injury events per video. The bleeding and bile spillage events do not represent separate injuries, but rather each occurrence of blood or bile spillage. These error frequencies are expected to be higher than what one would observe in a real procedure as binary classification thresholds are tuned to maximizing instance-level F2, which prioritizes recall without sacrificing too much precision.

**Table 2.** Performance of our error model on full-length videos for the three errors defined in the LC ontology. “Video-level” refers to metrics computed for all error instances in a single video and then averaged across all videos in the dataset. “Instance-level” refers to metrics computed for all instances across the entire dataset. We demonstrate high video-level and instance-level recalls and moderate video-level and instance-level precision.

| Error Type | Video-Level Recall (%) | Video-Level Precision (%) | Video-Level Accuracy of Error Presence (%) | Instance-Level Recall (%) | Instance-Level Precision (%) |
| --- | --- | --- | --- | --- | --- |
| Bleeding | 62.55 | 18.43 | 93.41 | 55.75 | 15.62 |
| Bile Spillage | 42.57 | 16.09 | 60.75 | 39.39 | 16.30 |
| Thermal Injury | 37.68 | 28.67 | 80.39 | 30.65 | 21.69 |

### Surgical Error Correlation Analysis

We analyze the error detection model’s outputs by observing how they correlate with known metadata, as shown in Figure 3. When examining the differences among errors by surgical step, Step 3 (Dissection of the Hepatocystic Triangle), the longest dissection step, had the highest total instances of bleeding (p < 0.01), while Step 2 (Exposure of the Gallbladder), the shortest dissection step, had the lowest total instances for all error types (p < 0.01). After normalizing for step duration, Step 3 continued to exhibit the highest bleeding instances rate (p < 0.01) but had the lowest rates of predicted bile spillage and thermal injury instances (p < 0.01).

Stratifying by disease severity, cases involving severe gallbladder disease (DSS 3 and 4) showed more total predicted error instances compared to cases with less severe disease (DSS 1 and 2) (p < 0.02). Similar to above, after normalizing for the duration of the entire active procedure (Steps 2 through 6), higher disease severity (DSS 3 and 4) was associated with increased rates of bleeding and bile spillage (p < 0.01), but a lower rate of thermal injury (p < 0.01).

**Figure 2.**
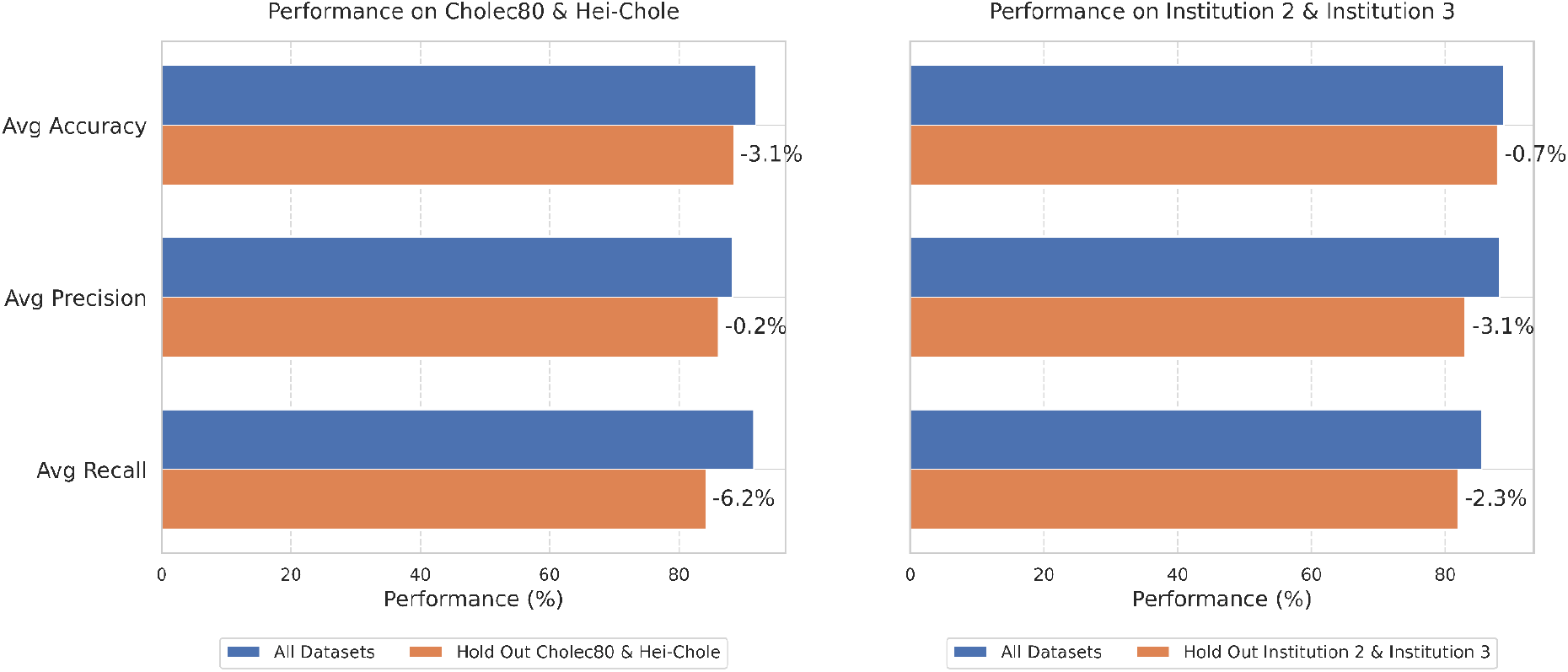
Bar charts that show the generalizability of our Surgical Step Model when evaluating on held-out datasets from training. “All Datasets” refers to training on the combined training splits of Cholec80, Hei-Chole, SST, Institution 1, Institution 2, and Institution 3. “Hold Out {datasets}” refers to training on the training splits of all datasets except those specified in {datasets}. We further assess the model’s ability to generalize to surgical videos from other hospitals, evaluating its performance on datasets (Table 1) held out from training. As shown in Figure 2 (left), when trained on Institutions 1, 2 and 3, and SST datasets and tested on Cholec80 and Hei-Chole, the model experienced only modest performance drops of 3.1% in accuracy, 0.2% in precision, and 6.2% in recall. Similarly, as shown in Figure 2 (right), when trained on the Institution 1, SST, Cholec80, and Hei-Chole datasets and tested on Institutions 2 and 3, the performance only drops 0.7% in accuracy, 3.1% in precision, and 2.3% in recall.

**Figure 3.**
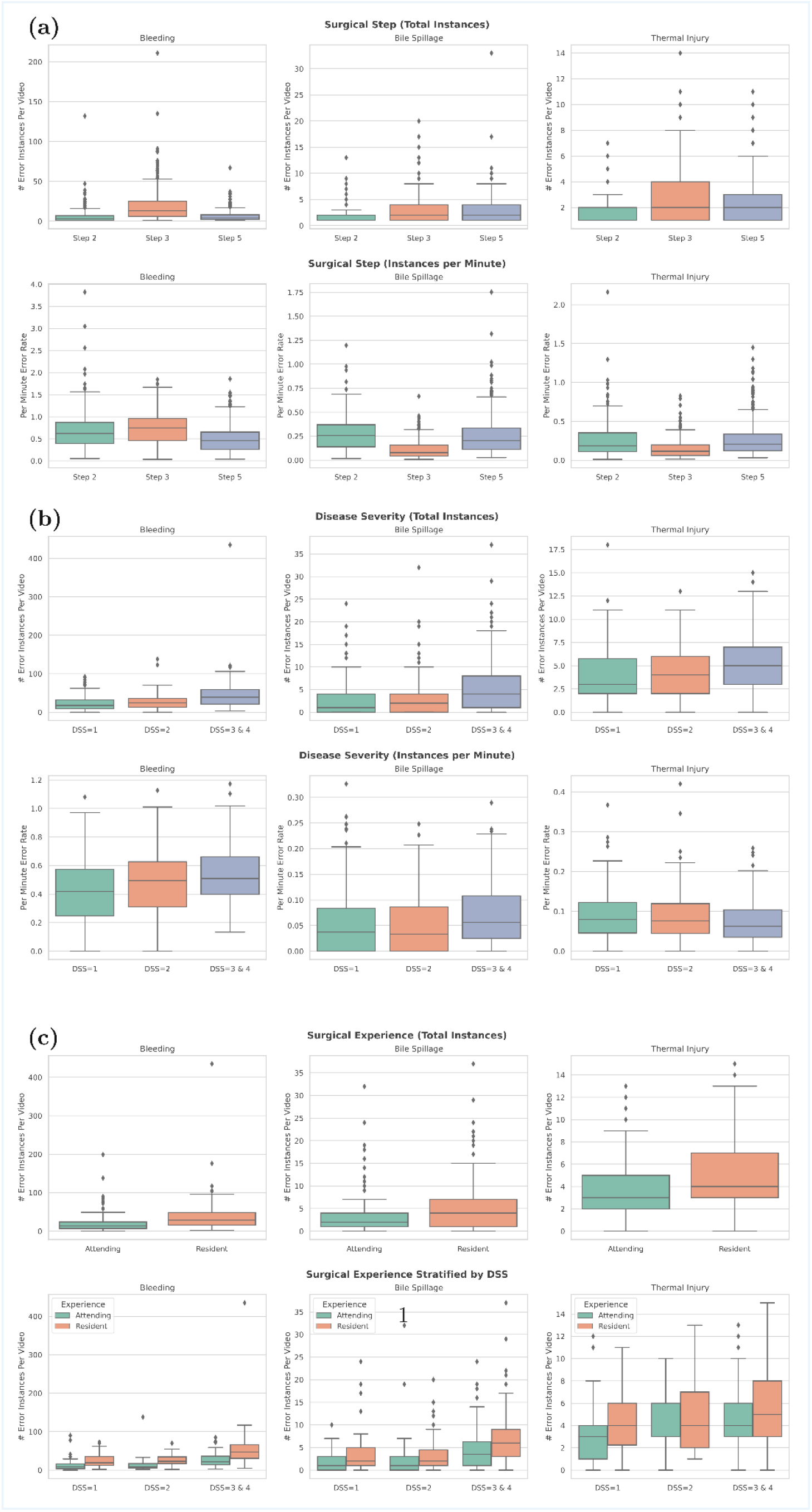
Box and whisker plots of the frequency and rate of predicted error instances for different categories of metadata. **a.***Top:* Distribution of per-video total number of predicted error instances for each dissection step over all videos. *Bottom:* Distribution of per-minute predicted error instance rates for each dissection step over all videos. Step 3 has the highest total instances of bleeding (p < 0.01), and step 2 has the lowest total predicted instances (p < 0.01). After normalizing for step duration, step 3 continues to have the highest bleeding prediction instance rate, but the lowest bile spillage and thermal injury rates (p < 0.01). Step 2 = Exposure of the gallbladder; Step 3 = Dissection of hepatocystic triangle; Step 5 = Gallbladder dissection. **b**. *Top:* Distribution of per-video total number of predicted error instances categorized by disease severity score (DSS). *Bottom:* Distribution of per-minute predicted error instances. Cases with higher DSS have more predicted error instances than cases with lower DSS (p < 0.02). After normalizing for video length, we find that a higher DSS is correlated with increased bleeding and bile spillage rates (p < 0.01), but lower thermal injury rates (p < 0.01). **c**. *Top:* Distribution of per-video total number of predicted error instances in cases performed by residents vs. attendings. *Bottom:* Distribution of per-video total number of predicted error instances in cases performed by residents vs. attendings stratified by DSS. We find that cases performed by residents have more predicted error instances overall than attendings (p < 0.01). Further stratified by disease severity, residents still showed higher incidence of all predicted error instances for lower diseased cases (p < 0.03), but only significantly more bleeding and bile errors (p < 0.02) for higher diseased cases.

Stratifying by surgeon skill level, cases performed by residents had a higher number of overall predicted error instances compared with cases performed by an attending surgeon (p < 0.01). This finding is the same when looking only at cases with no cholecystitis (DSS 1) (p < 0.03). Similarly, in cases with more severely diseased gallbladders (DSS 3 and 4), residents demonstrated a higher incidence of predicted bleeding and of bile spillage (p < 0.02).

## DISCUSSION

In this study, we developed an automated system to efficiently navigate and retrieve key segments of full-length laparoscopic cholecystectomy (LC) procedures at multiple temporal scales, focusing on surgical steps and errors. Using a LC-specific ontology comprising five high-level surgical steps and three fine-grained errors (bleeding, bile spillage, and thermal injury), we collected and annotated the largest multi-institution dataset to train Computer Vision (CV)-based models for this task. This dataset is over eight times larger than the current largest annotated LC dataset for surgical steps^10^ and over five times larger than the current largest annotated laparoscopic dataset for errors^19^ used in CV model training and evaluation.

Our step model, trained to classify the surgical step of each video frame, demonstrated strong performance and additionally showed strong generalization to surgical videos from new hospitals without significant performance loss. This study is the first to evaluate real-world generalizability for automated identification of surgical steps. Such generalizability is particularly valuable for hospitals that lack the resources to collect their own large datasets or to train in-house models, as they can deploy a model trained on videos from other hospitals to achieve reliable results.

Beyond CV-facilitated retrieval of surgical steps, we developed a model capable of retrieving fine-grained surgical errors. Our model was trained to localize the starting point of each predicted error instance, focusing on events significant in terms of tissue injury location and affected area size, as defined by trained general surgeons. These events are particularly relevant for distinguishing surgeon skill levels and are valuable for surgical training, performance assessment and feedback. We demonstrated that our error model performance is consistent with postoperative review settings, where high recall is prioritized to capture as many true error events as possible. Although we cannot guarantee that the model only flags clinically severe events, all detected events are brief, and true positives can be quickly and easily confirmed by a human reviewer. Precision is sufficient to keep false-positive filtering manageable, supporting efficient and effective error identification for review and analysis.

We further conduct a large-scale analysis of the predicted error instances, stratifying them by surgical step, disease severity, and surgeon experience level. We demonstrate that our findings are consistent with existing clinical knowledge. Specifically, we observe Step 3 having the highest rate of predicted bleeding which is unsurprising as it involves opening the gallbladder peritoneum and dissecting the hepatocystic triangle, where there is a high risk of bleeding, particularly in cases with higher disease severity. This step also has the lowest bile spillage and thermal injury rates which is likely due to the dissection’s proximity to vital structures such as the common bile duct and the duodenum, where dissection tends to be more careful to avoid injury. We further observed that Step 2, despite having the fewest error instances overall, exhibits a higher rate of bile spillage due to the inclusion of intentional gallbladder punctures in this step. Moreover, the higher spillage rate in Step 5 can likely be attributed to the higher degree of difficulty in differentiating between the proper dissection plane and the gallbladder parenchyma when removing it from the gallbladder fossa. Furthermore, bile spillage at this step is less critical than injury to the common bile duct.

We further observed that higher disease severity was associated with an increased number of predicted error instances, which is expected given the inflamed surgical field and also observed by Tranter-Entwistle, et al.^30^ Conversely, we observed a lower predicted instance rate of thermal injury in cases with a higher disease severity, as could be explained by a more careful dissection strategy used for more difficult cases. Moreover, we found a higher number of overall predicted error instances in cases performed by residents compared to those performed by an attending surgeon. This higher frequency likely reflects the residents’ yet-to-be-refined tissue handling and psychomotor skills. Overall, our findings are consistent with those derived from ground truth annotations, highlighting that our trained model aligns well with clinician assessments. While no unexpected correlations were found, our large-scale analysis demonstrates the potential of our model to detect both consistent and anomalous cases.

Traditionally, surgical training occurs in high-stakes operating rooms where surgical trainees are allowed to fully or partially perform a procedure under the supervision of an attending surgeon. These new CV-based models, though still a proof-of-concept, can already contribute to optimizing surgical training through quick navigation to key sections or moments of a procedure during postoperative surgical review. This can encourage trainees to curate their case videos for self-study and discussion with attending surgeons, thereby facilitating a faster ascent up their operative learning curves.

### Limitations

This study is limited to intracorporeal videos of LC, focusing on five surgical steps within the Execution of Objectives phase and three common errors. The framework we developed, however, can be readily extended to other surgical procedures, incorporating their unique steps and error types. Additionally, our error definitions do not distinguish between different levels of injury severity and our models do not factor in variables such as surgeon fatigue or case complexity, though in practice, these distinctions may significantly impact patient outcomes. A discrepancy was observed between the number of human-annotated errors and the model output. However, because the error segments are brief, we believe false positives can be easily identified and excluded during postoperative review. Future research could examine how to integrate these variables into a more comprehensive error ontology and modelling strategy, and expand model deployment to other laparoscopic procedures. Moreover, the metadata from our video sources do not include information on the number of cases the resident has performed, the level of attending supervision, or whether the attending surgeon took over. Therefore, the error correlation results with respect to surgical experience should be interpreted with caution. Finally, our analysis is restricted to postoperative video review. In the future, studies could investigate the potential application of these technologies in live surgical settings to prevent errors in real-time, especially those leading to life-threatening complications such as common bile duct injuries.

## Data Availability

Data could be shared upon reasonable request and following data sharing agreement between institutions. The code for training and running inference with the Computer Vision models will be made available upon publication.

## ACKNOWLEDGEMENTS

This study was funded by Wellcome Leap SAVE (No. 63447087-287892). The funder played no role in study design, data collection, analysis and interpretation of data, or the writing of this manuscript.

