## Supplemental for "Computer Vision-Based Retrieval of Steps and Errors in Laparoscopic Cholecystectomy"

**Supplemental Content 1: Annotation Ontology Details**

**Supplementary Table 1:** *Surgical Step Ontology*

**Supplementary Table 2:** *Disease Severity Grading Criteria*

**Supplemental Content 2: Model Architecture and Training Details**

**Supplementary Figure 1.** *Step Segmentation Model Architecture.*

**Supplementary Figure 2.** *Error Detection Model Training.*

**Supplementary Figure 3.** *Error Detection Inference Pipeline on Full-Length Videos.*

**Supplemental Content 3: Detailed Step Model Performance Results**

**Supplementary Table 3.** *Step model performance on all datasets.*

**Supplemental Content 4: Detailed Error Model Performance Results**

**Supplementary Table 4.** *Framewise binary classification performance on blood, bile and thermal injury burn detection.*

**Supplementary Table 5.** *Performance of our state-change detection model for different error types, temporal sampling rates and randomization strategies.*

**Supplemental Content 5: Surgical Error Correlation Analysis Method**

**Supplemental Content 6: Correlation Analysis on Ground Truth Annotations**

**Supplementary Figure 4.** *Frequency and Rate of Ground Truth Error Instances For Different Categories of Metadata.*

### Supplemental Content 1: Annotation Ontology Details

**Supplementary Table 1.** Detailed ontology of intracorporeal steps in the Execution of Surgical Objectives phase of laparoscopic cholecystectomy.

| | **2 Expose the Gallbladder** | **3 Dissection of Gallbladder / Hepatocystic triangle** | **4 Ligation & Division** | **5 Gallbladder Dissection** | **6 Packaging & Removal** |
|---|---|---|---|---|---|
| **Start frame** | Last port is inserted | First dissection of the gallbladder peritoneum | Instrument enters with the intention to ligate the Cystic Artery or the Cystic Duct | Both structures are divided and the instrument is out of view | The gallbladder leaves the liver bed |
| **End frame** | First dissection of the gallbladder peritoneum | Instrument enters with the intention to ligate the Cystic Artery or the Cystic Duct | Both structures are divided and the instrument is out of view | The gallbladder leaves the liver bed | The gallbladder leaves the intra abdominal cavity |

**Supplementary Table 2.** *Grading criteria for Disease Severity Score (DSS).*

| **Score** | **Criteria** |
|---|---|
| 1 | Thin-walled, normal-appearing gallbladder with no adhesions |
| 2 | Mildly abnormal-appearing gallbladder (slightly thick-walled or distended) and/or has thin, filmy adhesions |
| 3 | Moderately abnormal-appearing gallbladder (thick-walled, with mucocele, or large distended gallbladder) and/or has overlying moderate adhesions |
| 4 | Severely inflamed or grossly abnormal-appearing gallbladder (e.g. necrotic or perforated) and/or has extensive or dense adhesions |

### Supplemental Content 2: Model Architecture and Training Details

#### Step Recognition Model

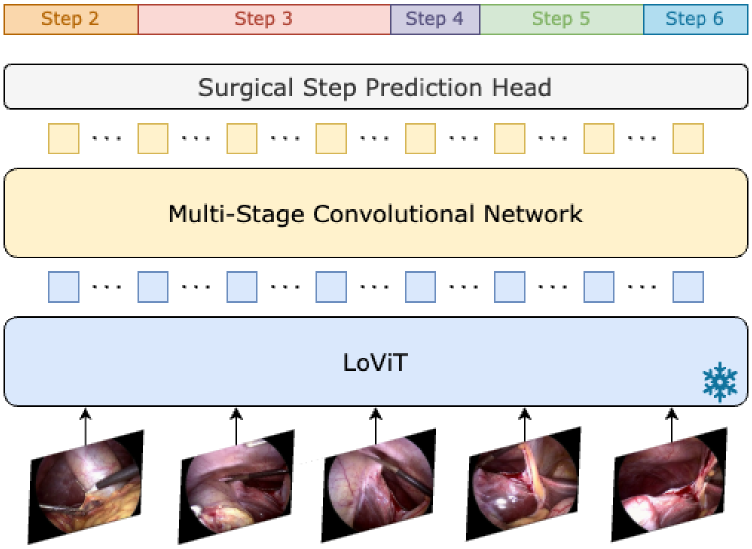


**Supplementary Figure 1. Step Segmentation Model Architecture.**

To segment the steps of a LC, we extracted LC-relevant visual features for frames sampled at 1 fps using the LoViT[1] encoder. LoViT is a Vision Transformer-based[2] model that is trained on LC frames from Cholec80[3] and AutoLaparo[4]. We trained Multi-Stage Convolutional Network (MS-TCN)[4] architecture adapted for framewise classification of a video for longer durations (LTContext[5]). Details of this model are provided in Supplementary Figure 1.

#### Error Detection Model

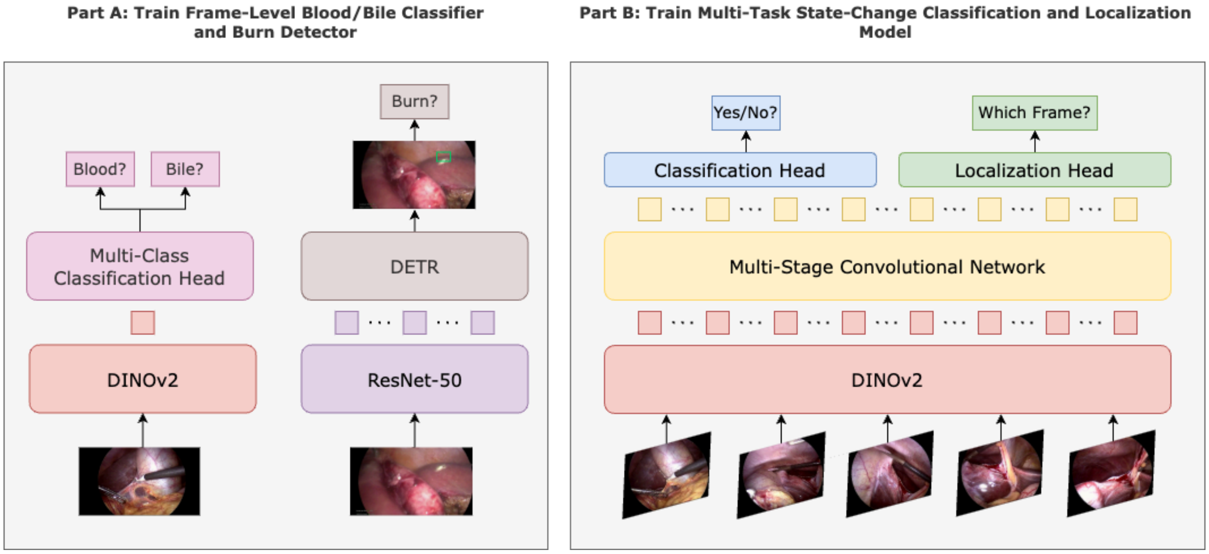


**Supplementary Figure 2. Error Detection Model Training.** Training consists of two parts: A) training a frame-wise classification or detection model for blood/bile/burn classification. B) training a state-change detection model to classify whether a new injury occurs in a short clip.

We propose a two-stage pipeline for intraoperative error detection, 1) frame-based binary classification to detect the presence or absence of the result of injury (blood, bile, thermal injury), noting state changes where the detection changes from absence to presence, and 2) clip-based binary classification of whether a state change occurred at sampled clips around the detected points and within each continuous segment of a detected injury result (blood/bile/thermal injury).

To train the frame-based binary classification model, we created a dataset of randomly sampled frames from our video dataset with more dense sampling around identified error occurrences. As blood and bile can be easily identified in a frame, we utilize an image classification model to classify the presence of blood and bile in each frame. Blood was annotated to be present in a frame if it includes blood that is fresh, pooled, pouring, coagulated, and/or staining, but does not include uninjured vessels. Bile was annotated to be present in a frame if it includes bile or bile stones. Our image classification model builds off the pre-trained vision foundation model DINOv2[6]. Given its pre-training on 142 million general images, it has a rich understanding of visual features. As depicted in Supplementary Figure 2 (left), we adapt this model by simply adding a linear layer on top of the DINOv2 backbone and fine-tuning the entire model with Low-Rank Adaptation (LoRA)[7] to preserve the backbone's learned visual understanding from pre-training and effectively adapting it to our smaller LC dataset. Since thermal injuries result in very fine-grained, but visually defined, burn marks, we utilize an object detection model to identify its spatial location. We then collapse the spatial labels into framewise predictions. Burn marks were annotated as any burned area where due to thermal injury to tissue outside of the dissection area. As depicted in Supplementary Figure 2 (left), we use the Deformable DETR model[8], as the marks are very spatially small yet visually distinct. We fine-tuned it from the MS COCO[9] pretrained checkpoint.

To train the state-change binary classification model, we created a dataset of sampled 16-frame clips from our video dataset where 20% of clips contain a state change due to the occurrence of an error and the remaining do not. Given that these errors do not occur frequently within an entire procedure, we attempt to mimic the sparseness of the occurrences with our data split. As shown in Supplementary Figure 2 (right), we build off the rich DINOv2 backbone by adding an MS-TCN to model the temporal dependencies within each video clip and two linear classification heads on top of the pooled temporal features to determine the occurrence of a state change and localize where the state change occurs within the clip. We fine-tune the entire model with LoRA.

#### Full Error Detection Pipeline

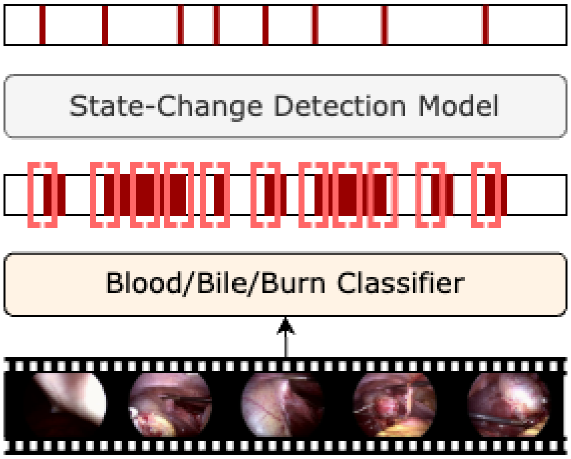


**Supplementary Figure 3. Error Detection Inference Pipeline on Full-Length Videos.** Inference consists of 3 steps: 1) Sampling frames at 10 fps and performing frame-wise classification for blood/bile/burn presence. 2) Extracting candidate segments for state change. 3) Classifying the candidate segments using the state-change detection model to remove false positives.

As shown in Supplementary Figure 3, to perform error detection on a full-length video, frames are first extracted at 10 fps and frame-wise classification is performed. Candidate clips of 16-frames at different temporal resolutions (1, 2, 5, 10 fps) are sampled around where the frame-wise classification changes from absence to presence and throughout a continuous segment of presence above a certain threshold. For each frame rate, state change detection models trained for that particular temporal resolution are applied to the corresponding candidate clips. Predictions over all state change models are aggregated and predictions over 0.4 confidence are kept. Thresholds for classification of each error type are chosen such that the instance-level F2 is maximized on the validation set over a grid of thresholds between 0.4 and 1.0 at intervals of 0.05. We choose to maximize F2 as recall is favoured over precision given that one would like to capture as many examples as possible and filtering false positives is very quick.

#### Supplemental Content 3: Detailed Step Model Performance Results

**Supplementary Table 3. Step model performance on all datasets. Average metrics are reported over all videos in the dataset(s).**

| **Dataset** | **Average Accuracy (%)** | **Average Recall (%)** | **Average Precision (%)** |
|---|---|---|---|
| Cholec80 | 93.0 | 93.1 | 89.6 |
| Hei-Chole | 87.9 | 84.1 | 84.0 |
| SST | 85.7 | 84.8 | 79.7 |
| Institution 1 | 88.9 | 87.7 | 84.3 |
| Institution 2 | 91.9 | 88.7 | 91.8 |
| Institution 3 | 85.5 | 78.3 | 79.6 |
| Average | 88.3 | 86.6 | 83.1 |

#### Supplemental Content 4: Detailed Error Model Performance Results

##### Framewise Classification

To identify the presence of blood and bile in a frame, we create a dataset of 26,404 images where 24,551 images are annotated with the presence/absence of blood and 9,843 images are annotated with the presence/absence of bile. Of the annotated images for blood, 73.9% contained blood and of the annotated images for bile, 45.4% contained bile. As blood and bile are fairly visually similar, we train a multiclass blood and bile classification model so that the model may learn how to differentiate between the two. As thermal injury burns are visually distinct but are generally small in size, we instead train an object detection model on a dataset of 4,743 images annotated with thermal injury spatial annotations, where a total of 8,174 instances were annotated. We further post-process the detections into framewise classifications for use in our error detection pipeline. All images were sampled from videos in our dataset and models were trained according to the data split used for error detection. Supplementary Table 4 illustrates the classification performance of our classification models.

**Supplementary Table 4. Framewise binary classification performance on blood, bile and thermal injury burn detection.**

| **Error Type** | **Accuracy (%)** | **AUROC (%)** |
|---|---|---|
| Blood | 89.16 | 95.10 |
| Bile | 76.73 | 87.24 |
| Thermal Injury Burn | 81.40 | 84.88 |

#### State Change Detection

To identify whether a state change occurs in a short clip, we create a dataset of 16-frame clips sampled from videos in our dataset such that approximately 20% of the clips in the training set contain a state change and 80% do not. As error occurrences are relatively sparse in full-length procedures, we trained the model on a dataset that mimicked this sparsity. As we do not want our model to be biased towards a particular temporal position of state change, we sample clips such that the location of the state change is randomly selected according to a Gaussian distribution around the middle frame or a Uniform distribution over the entire clip. We follow the same data split used for error detection. Overall, we sampled 6,431-6,490 clips for bleeding state change, 4,675-5,282 clips for bile spillage state change, and 5,244-5,323 clips for thermal injury state change. Supplementary Table 5 reports the binary classification performance for each state change model.

**Supplementary Table 5. Performance of our state-change detection model for different error types, temporal sampling rates and randomization strategies.**

| **Error** | **Sampling rate (fps)** | **Randomization Strategy** | **AUROC (%)** |
|---|---|---|---|
| Bleeding | 5 | Gaussian | 70.00 |
| | 5 | Uniform | 63.49 |
| | 2 | Gaussian | 70.42 |
| | 2 | Uniform | 69.85 |
| | 1 | Gaussian | 74.46 |
| | 1 | Uniform | 68.53 |
| Bile Spillage | 5 | Gaussian | 76.40 |
| | 5 | Uniform | 76.11 |
| | 2 | Gaussian | 75.62 |
| | 2 | Uniform | 73.01 |
| | 1 | Gaussian | 74.51 |
| | 1 | Uniform | 73.17 |
| Thermal Injury | 10 | Gaussian | 87.37 |
| | 10 | Uniform | 82.86 |
| | 5 | Gaussian | 81.37 |
| | 5 | Uniform | 75.93 |
| | 2 | Gaussian | 80.75 |
| | 2 | Uniform | 79.61 |

#### Supplemental Content 5: Surgical Error Correlation Analysis Method

We used the Mann–Whitney U test[10] and a Kruskal–Wallis test[11] to identify potential correlations between error frequency and step, disease severity score, surgical experience (attending vs. resident), and data source. Both the Mann–Whitney U and Kruskal–Wallis tests are non-parametric statistical tests used to compare two and three or more independent groups. For categories with three or more independent groups, we first apply the Kruskal–Wallis test to determine if there are any significant differences among the groups and if so, apply the Mann–Whitney U test on all permutations of pairs of subgroups to determine if one group is significantly greater than the other. As we are performing multiple statistical tests, we use Bonferroni correction[12] to adjust the default p-value of 0.05 for significance.

#### Supplemental Content 6: Correlation Analysis on Ground Truth Annotations

We also performed a large-scale analysis of our video dataset with respect to ground truth error annotations, as illustrated in Supplementary Figure 4. We found that most significant findings observed using the model predictions are the same as those found with ground truth. The only differences were that using the ground truth annotations, there were no significant differences in the total number of bile spillage instances across steps, and there were no significant differences between the bleeding error rates per minute across steps. Furthermore, there were no significant differences comparing error instances of bile spillage and thermal injury for residents and attendings. However, when comparing the median values of error instances across the different groups, we observe either a positive trend or no clear trend for significant correlations derived from model predictions that were insignificant when derived from ground truth.

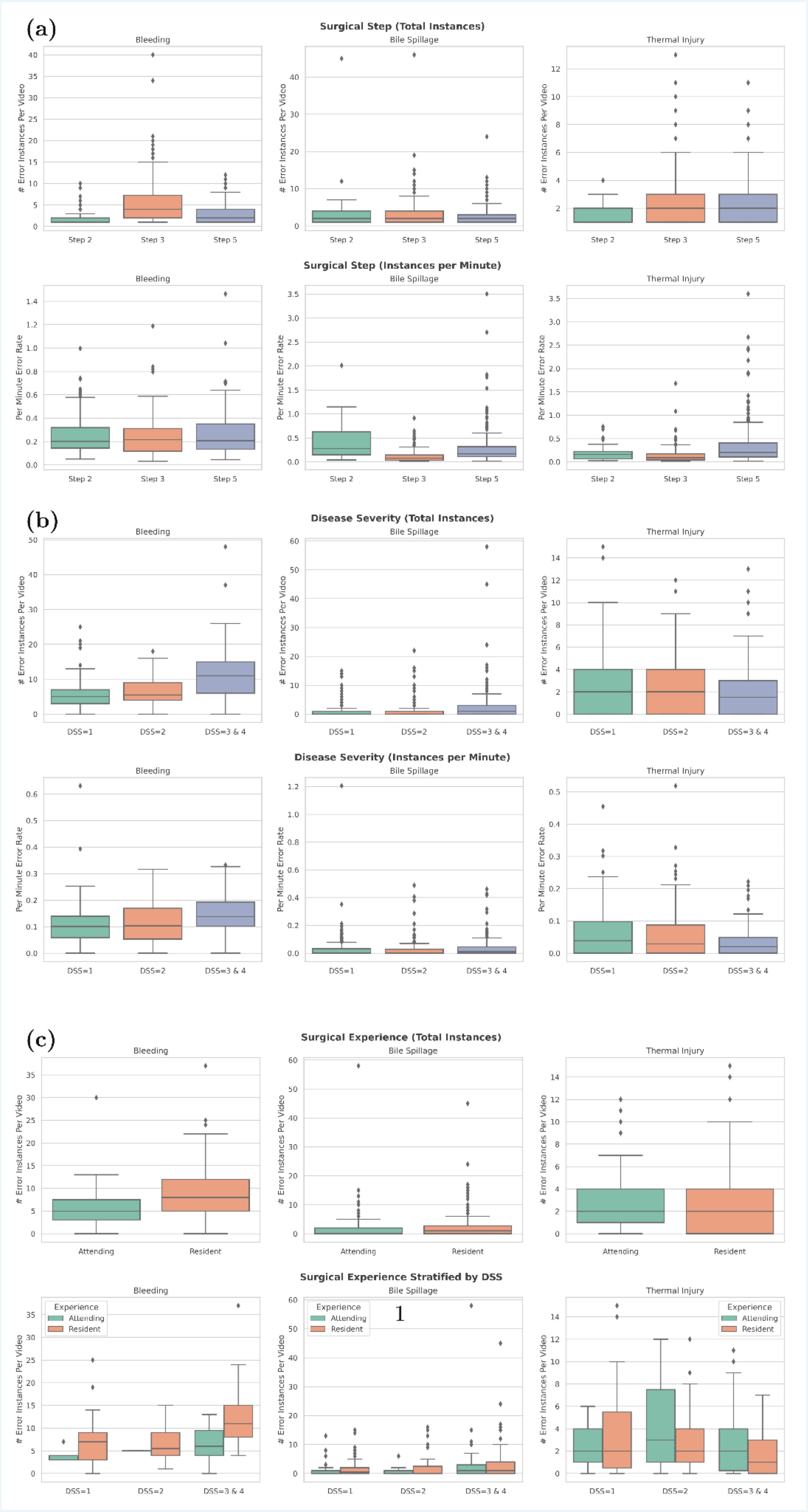
(a)
Surgical Step (Total Instances)
Bleeding
Bile Spillage
Thermal Injury
### Error Instances Per Video
Step 2
Step 3
Step 5
Surgical Step (Instances per Minute)
Per Minute Error Rate
(b)
Disease Severity (Total Instances)
DSS=1
DSS=2
DSS=3 & 4
Disease Severity (Instances per Minute)
(c)
Surgical Experience (Total Instances)
Attending
Resident
Surgical Experience Stratified by DSS
Experience
Attending
Resident
1

**Supplementary Figure 4. Frequency and Rate of Ground Truth Error Instances For Different Categories of Metadata. a.** *Top:* Distribution of per-video total number of error instances for each dissection step over all videos. *Bottom:* Distribution of per-minute error rates for each dissection step over all videos. We find that step 3 has the highest total instances of bleeding ($p < 0.01$), and step 2 has the lowest total bleeding and thermal injury error instances ($p < 0.01$). After normalizing for step duration, we find that step 2 has the highest bile spillage rate ($p < 0.01$), and step 5 has the highest thermal injury rate ($p < 0.04$), and step 3 has the lowest bile spillage and thermal injury rates ($p < 0.01$). **b.** *Top:* Distribution of per-video total number of error instances for each disease severity score over all videos with annotated disease severity. *Bottom:* Distribution of per-minute error rates for each disease severity score over all videos with annotated disease severity. We find that cases with higher diseased gallbladders have more bleeding and bile spillage instances than lower diseased ones ($p < 0.01$). After normalizing for video length, we find that higher diseased cases are correlated with increased bleeding and bile spillage rates ($p < 0.01$), but lower thermal injury rates ($p < 0.01$). **c.** *Top:* Distribution of per-video total number of error instances in cases performed by residents vs. attendings for cases with annotated surgical experience. *Bottom:* Distribution of per-video total number of error instances in cases performed by residents vs. attendings stratified by disease severity score. We find that cases performed by residents have more bleeding errors than those performed by attendings ($p < 0.02$). Further stratified by disease severity, residents still showed significantly higher incidence of bleeding for high disease cases ($p < 0.01$).
